# Effects of Sacubitril/valsartan on Hypertensive Heart Disease: the REVERSE-LVH Randomized Phase 2 Trial

**DOI:** 10.1101/2025.04.08.25325450

**Authors:** Vivian Lee, Mayank Dalakoti, Qishi Zheng, Desiree-Faye Toh, Redha Boubertakh, Jennifer A Bryant, Tar-Choon Aw, Chi-Hang Lee, A Mark Richards, Javed Butler, Javier Díez, Roger Foo, Stuart A Cook, Carolyn SP Lam, Thu-Thao Le, Calvin WL Chin

**Affiliations:** National Heart Research Institute of Singapore, National Heart Centre Singapore, Singapore, Singapore; Department of Cardiology, National University Heart Centre Singapore, National University Health System, Singapore, Singapore; Cochrane Singapore, Singapore, Singapore; Department of Cardiology, National Heart Centre Singapore, Singapore, Singapore; Department of Laboratory Medicine, Changi General Hospital, Singapore, Singapore; Christchurch Heart Institute, University of Otago, Christchurch, New Zealand; Baylor Scott and White Research Institute, Dallas, Texas, United States; Department of Medicine, University of Mississippi School of Medicine, Mississippi, United States; Centre of Applied Medical Research (CIMA) and School of Medicine, University of Navarra, Pamplona, Spain; Centre for Network Biomedical Research of Cardiovascular Diseases (CIBERCV), Carlos III Institute of Health, Madrid, Spain; Cardiovascular Metabolic Disease Translational Research Program, Yong Loo Lin School of Medicine, National University of Singapore, Singapore, Singapore; Cardiovascular & Metabolic Disorders SRP, Duke-NUS Medical School, Singapore, Singapore; Cardiovascular Medicine Academic Clinical Program (ACP), Duke-NUS Medical School, Singapore, Singapore

**Keywords:** Diffuse interstitial fibrosis, cardiovascular magnetic resonance, hypertensive heart disease

## Abstract

Diffuse interstitial fibrosis is associated with adverse outcomes in hypertensive heart disease and may be reversible. Sacubitril/valsartan could offer greater anti-fibrotic effects than valsartan alone. In the REVERSE-LVH phase 2 open-labelled trial (clinicaltrials.gov NCT: 03553810; funded by the National Medical Research Council of Singapore), 78 patients with essential hypertension and left ventricular hypertrophy (LVH) were randomized 1:1 to sacubitril/valsartan or valsartan for 52 weeks. Primary endpoint was a change in interstitial volume, assessed using cardiovascular magnetic resonance. Despite similar 24-hour systolic blood pressure at 52 weeks (125±11 vs. 126±11 mmHg; P=0.379), sacubitril/valsartan resulted in a greater absolute reduction in interstitial volume compared to valsartan (−5.2±5.4 vs. −2.5±3.1 mL; P=0.006). Secondary endpoints showed significant differences favoring sacubitril/valsartan in LV mass, left atrial volume, estimated LV filling pressure, and improved cardiac circulating biomarkers (N-terminal pro-B-type natriuretic peptide and high-sensitivity troponin T). Other markers of cardiac volumes, function and mechanics were similar between the two treatment arms. Here we show the potential myocardial benefits of sacubitril/valsartan beyond blood pressure control, though larger studies are needed to confirm their clinical relevance.

## INTRODUCTION

About 30% of the burden related to hypertension occurred in individuals with well-controlled blood pressures (BP)^1^. The updated definition of heart failure (HF) stages had identified more asymptomatic individuals in a pre-clinical stage^2^. This suggests that despite substantial benefits associated with BP lowering, there remains opportunities to improve risk stratification and develop therapeutic options beyond managing peripheral BP to slow/prevent the development of symptomatic HF.

In hypertensive heat disease (HHD), left ventricular hypertrophy (LVH) occurs in response to hemodynamic and neuro-hormonal stresses triggered by prolonged exposure to elevated BP^3^. Its pathophysiology is accompanied by an expansion in extracellular volume (ECV) due to accumulation of extracellular matrix components and collagen deposition throughout the myocardium – diffuse interstitial fibrosis. This type of fibrosis typically develops gradually and is initially reversible^4^. Myocardial fibrosis significantly contributes to cardiac dysfunction, arrhythmias, and reduced coronary perfusion, ultimately leading to HF and adverse cardiovascular outcomes^4,5^. Therefore in addition to peripheral BP, diffuse interstitial fibrosis is a potential marker for enhancing risk stratification and directing targeted therapies to prevent the progression of hypertensive LVH to advanced HF stages.

Angiotensin receptor–neprilysin inhibitor (ARNI) represents a novel drug class that inhibits the renin-angiotensin-aldosterone system (RAAS) and enhances natriuretic peptides. This dual agent combines a neprilysin inhibitor (sacubitril) and an angiotensin receptor blocker (ARB; valsartan). Sacubitril/valsartan has shown superior benefits over conventional ARB or angiotensin-converting enzyme inhibitors (ACEI) monotherapy in lowering BP and improving outcomes in those with HF and reduced LV ejection fraction (HFrEF)^6,7^. Pre-clinical studies of mouse models have demonstrated that sacubitril/valsartan is effective in reducing myocardial fibrosis and improving cardiac function by attenuating angiotensin II and transforming growth factor b1 fibrotic pathways^8,9^.

Using cardiovascular magnetic resonance (CMR) to quantify interstitial volume as a measure of diffuse interstitial fibrosis non-invasively^10^, the aim of the study was to compare sacubitril/valsartan versus valsartan in reducing diffuse interstitial fibrosis in patients with hypertensive LVH. The hypothesis was that 52 weeks of sacubitril/valsartan therapy would result in greater regression of diffuse interstitial fibrosis compared to valsartan, independent of BP control.

## RESULTS

### Study Participants

A total of 80 patients were recruited, and 78 individuals were successfully randomized to sacubitril/valsartan (n=39) and valsartan (n=39) (Figure 1). The mean age at baseline was 58±11 years, and 32 (41%) were males. The baseline 24h mean SBP and DBP were 137±14 and 81±11 mmHg, respectively. The baseline clinical, renal function and CMR characteristics were well-balanced between the two treatment groups (Table 1). There was no change in body weight at the start and end of study.

**Figure 1.**
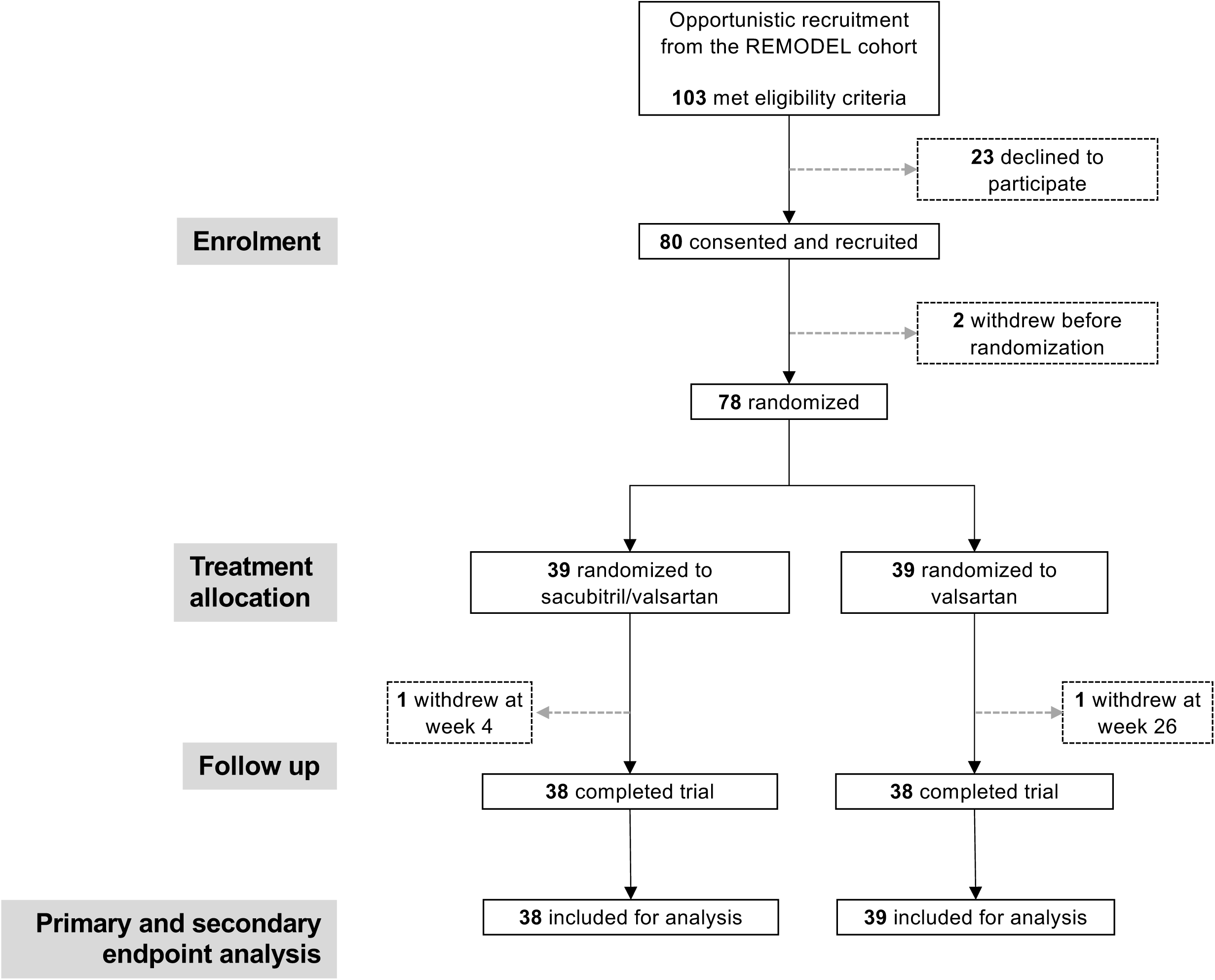
Participant Recruitment, Randomization and Treatment. A total of 78 participants were randomized in the trial. One patient in the sacubitril/valsartan group withdrew at week 4 due to concerns of medication side effects. One patient in the valsartan group withdrew at week 26 due to difficulties with trial commitment. This participant had final visit investigations (including cardiovascular magnetic resonance) performed at time of withdrawal, according to study protocol. Primary and secondary endpoints were analyzed as intention-to-treat.

**Table 1.** Baseline Characteristics of All Randomized Participants.

|  | All<br>Participants<br>(n=78) | Sacubitril/<br>valsartan<br>(n=39) | Valsartan<br>(n=39) |
| --- | --- | --- | --- |
| **Baseline Clinical Characteristics** |  |  |  |
| Age, y | 58±11 | 56±11 | 59±11 |
| Males, n (%) | 32 (41) | 18 (46) | 14 (36) |
| Chinese ethnicity, n (%) | 71 (91) | 35 (90) | 36 (92) |
| Weight, kg | 71±18 | 74±19 | 68±16 |
| Body mass index, kg/m<sup>2</sup> | 27.2±5.6 | 27.7±6.2 | 26.7±5.1 |
| Dyslipidemia, n (%) | 33 (42) | 18 (46) | 15 (38) |
| Diabetes mellitus, n (%) | 14 (18) | 9 (23) | 5 (13) |
| Smoking, n (%) | 6 (8) | 3 (8) | 3 (8) |
| 24h systolic blood pressure, mmHg | 137±14 | 138±17 | 137±10 |
| 24h diastolic blood pressure, mmHg | 81±11 | 82±11 | 80±11 |
| **Baseline Biochemical Characteristics** |  |  |  |
| Potassium, mmol/L | 3.8 [3.6, 4.0] | 3.8 [3.6, 4.0] | 3.7 [3.6, 4.0] |
| Creatinine, μmol/L | 72 [52, 86] | 73 [51, 88] | 72 [54, 85] |
| NT-proBNP, pg/mL | 53 [28, 141] | 42 [28, 101] | 60 [31, 275] |
| High sensitivity cTnT, pg/mL | 9.8 [6.5, 14.5] | 9.5 [6.6, 15.0] | 10.9 [6.0, 14.5] |
| Estimated GFR, mL/min/1.73 m<sup>2a</sup> | 90 [72, 106] | 94 [74, 108] | 88 [70, 102] |
| **Baseline Cardiovascular Magnetic Resonance Characteristics** |  |  |  |
| Indexed LV mass, g/m<sup>2</sup> | 64±19 | 64±19 | 64±18 |
| Indexed LV end diastolic volume, mL/m<sup>2</sup> | 78±15 | 76±16 | 79±15 |
| Indexed LV end systolic volume, mL/m<sup>2</sup> | 32±12 | 32±13 | 32±11 |
| Indexed LV stroke volume, mL/m<sup>2</sup> | 46±7 | 45±8 | 47±7 |
| LV ejection fraction | 60±9 | 59±10 | 60±7 |
| Indexed RV end diastolic volume, mL/m<sup>2</sup> | 74±13 | 73±13 | 74±13 |
| Indexed RV end systolic volume, mL/m<sup>2</sup> | 28±10 | 28±9 | 27±11 |
| Indexed RV stroke volume, mL/m<sup>2</sup> | 46±7 | 45±8 | 47±7 |
| RV ejection fraction | 63±9 | 62±8 | 64±9 |
| Indexed left atrial volume, mL/m<sup>2</sup> | 55±15 | 53±12 | 56±18 |
| Indexed right atrial area, cm<sup>2</sup>/m<sup>2</sup> | 12.0±2.1 | 11.7±1.8 | 12.4±2.2 |
| Remodeling Index | 5.33±0.99 | 5.22±0.80 | 5.44±1.15 |
| Ventricular-arterial coupling ratio | 0.71±0.29 | 0.74±0.36 | 0.69±0.22 |
| Estimated LV filling pressure, mmHg | 15.6±2.8 | 15.7±3.0 | 15.5±2.6 |
| Maximal wall thickness, mm | 10.0±2.4 | 10.1±2.1 | 9.9±2.7 |
| Mean wall thickness, mm | 7.5±1.6 | 7.7±1.6 | 7.4±1.6 |
| LV mass/end-diastolic volume ratio | 0.83±0.17 | 0.84±0.14 | 0.81±0.19 |
| Global longitudinal strain, % | -16.5±3.3 | -16.0±3.2 | -17.0±3.2 |
| Global circumferential strain, % | -18.7±3.2 | -18.2±3.5 | -19.1±2.8 |
| Global radial strain, % | 32.4±8.6 | 31.2±9.2 | 33.6±7.8 |
| Extracellular volume fraction, % | 26.4±2.6 | 26.1±2.4 | 26.8±2.8 |
| Indexed myocyte volume, mL/m<sup>2</sup> | 45.0±13.1 | 45.4±13.7 | 44.5±12.6 |
| Non ischemic late gadolinium enhancement, n (%) | 29 (37) | 15 (38) | 14 (36) |

Values were presented as mean ± standard deviations or median [interquartile range] unless otherwise indicated. <sup>a</sup>Estimated GFR calculated using MDRD equation = 175 x [serum creatinine (μmol/L)/88.4]<sup>-1.154</sup> × (age)<sup>-0.203</sup> × (0.742 if female). Left ventricular hypertrophy was defined based on age- and sex-specific reference ranges established in the local population (females: 47g/m<sup>2</sup> and males: 65g/m<sup>2</sup>). Abnormal global longitudinal strain in the population was defined as < -14.0 to -15.0% in males and < -17.0 to -18.0% in females<sup>20</sup>.
GFR = glomerular filtration rate; LV = left ventricular; RV = right ventricular; NT-proBNP = N-terminal pro-B-type natriuretic peptide; cTnT = cardiac troponin T

The median dose prescribed in the sacubitril/valsartan group was 200 [100, 200] mg twice a day and 160 [160, 320] mg once a day in the valsartan group, respectively. A total of 50 participants required additional anti-hypertensive medications (sacubitril/valsartan group, n=26 and valsartan group, n=24; P=0.571; Supplemental Table 2). The overall treatment adherence rate achieved in the trial was 97% (sacubitril/valsartan group, 97%; valsartan group, 96%).

### Primary Trial Endpoint

The baseline interstitial volume in the sacubitril/valsartan group was 29.2±12.8 mL compared to 28.0±11.3 mL in the valsartan group. After 52 weeks, sacubitril/valsartan resulted in greater regression in diffuse interstitial fibrosis compared to valsartan alone (-5.2±5.4 versus -2.5±3.1 mL, P=0.006), with an absolute between-group difference of -2.8 (95% CI: -4.8 to - 0.8) mL. Sacubitril/valsartan resulted in an 18% reduction in interstitial volume from baseline, compared to an 8.9% reduction with valsartan. The reduction in diffuse interstitial fibrosis remained greater in the sacubitril/valsartan group compared to valsartan alone after accounting for baseline interstitial volume, prior ACEI or ARB use, and baseline 24h SBP (Figure 2).

**Figure 2.**
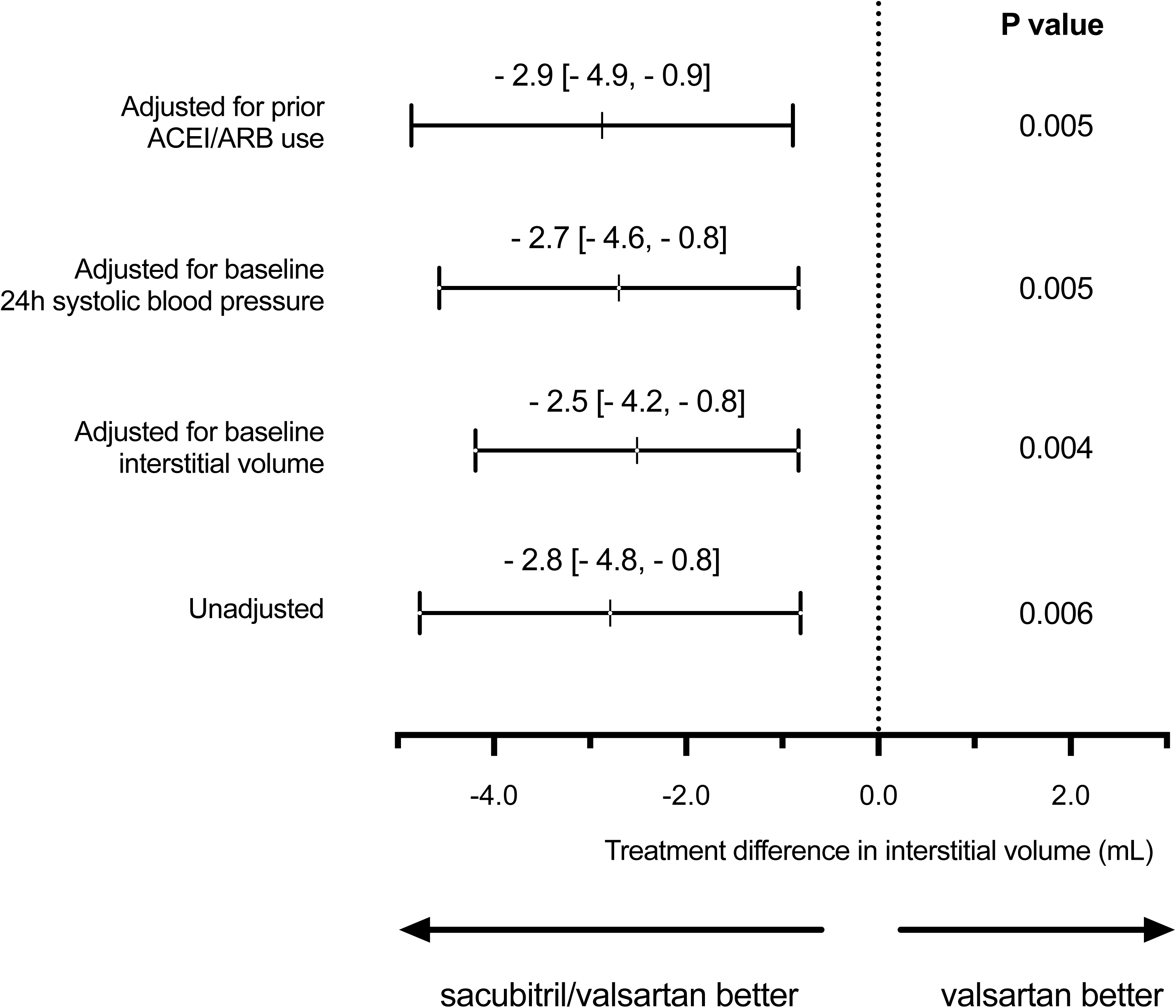
Effects of Sacubitril/valsartan Compared to Valsartan on Primary Endpoint. The primary endpoint in 78 participants. The effect estimates were presented in mean and 95% confidence intervals, with corresponding P values from one-way ANCOVA testing before and after adjustment for baseline covariates. Abbreviations: ACEI angiotensin-converting enzyme inhibitor; ARB, angiotensin receptor blocker.

Exploratory subgroup analysis suggested treatment heterogeneity in favor of sacubitril/valsartan in individuals with 24h median SBP >135 mmHg, with an absolute between-group difference of -5.2 (95% CI: -8.0 to -2.5) mL compared to -0.5 (95% CI: -3.2 to 2.2) mL in those with 24h SBP ≤135 mmHg (P=0.016 for interaction). There was no significant treatment heterogeneity in the other subgroups stratified by age, sex, LV ejection fraction, baseline interstitial volume, prior ACEI or ARB use and body mass index (Supplemental Table 3).

### Secondary Trial Endpoints

Compared to valsartan alone, sacubitril/valsartan resulted in greater reduction in LV mass (-21.9±25.4 versus -11.0±9.0 g, P=0.014) and myocyte volume (-16.0±19.5 versus -8.0±7.1 mL, P=0.019). Sacubitril/valsartan also led to a greater decrease in LA volume (-18.3±16.6 versus -8.6±13.6 mL, P=0.006) and LV filling pressures (-1.8±1.5 versus -0.9±1.1 mmHg, P=0.003) compared to valsartan, suggestive of improved diastolic function.

There were no significant differences in changes in cardiac volumes, systolic function, and other measures of myocardial mechanics between the treatment groups. Although sacubitril/valsartan was associated with a statistically significant improvement in global longitudinal strain (GLS) compared to valsartan (-0.9±2.3 versus 0.2±1.9, P=0.027), the effect size was small and the isolated improvement in GLS was not associated with improvement in the other strain components (Table 2). Similar findings were observed when relevant CMR metrics were indexed to body surface area (Supplemental Table 4). Compared to valsartan, sacubitril/valsartan was associated with a significant reduction from baseline to 52 weeks in NT-proBNP (ETR=0.72; 95% CI: 0.54–0.91; P=0.006) and hsTnT (ETR=0.83; 95% CI: 0.69–0.99; P=0.005).

**Table 2.**
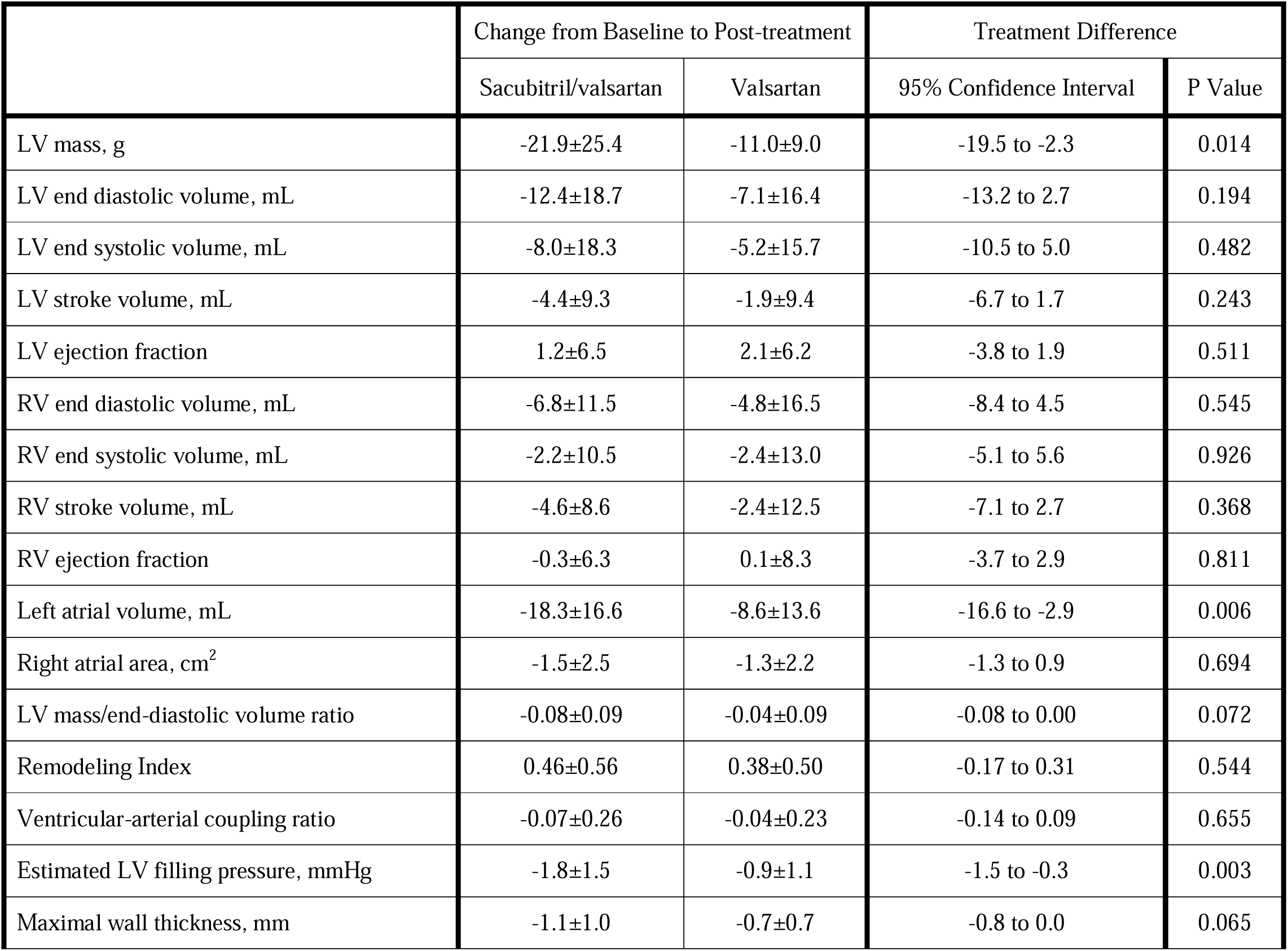
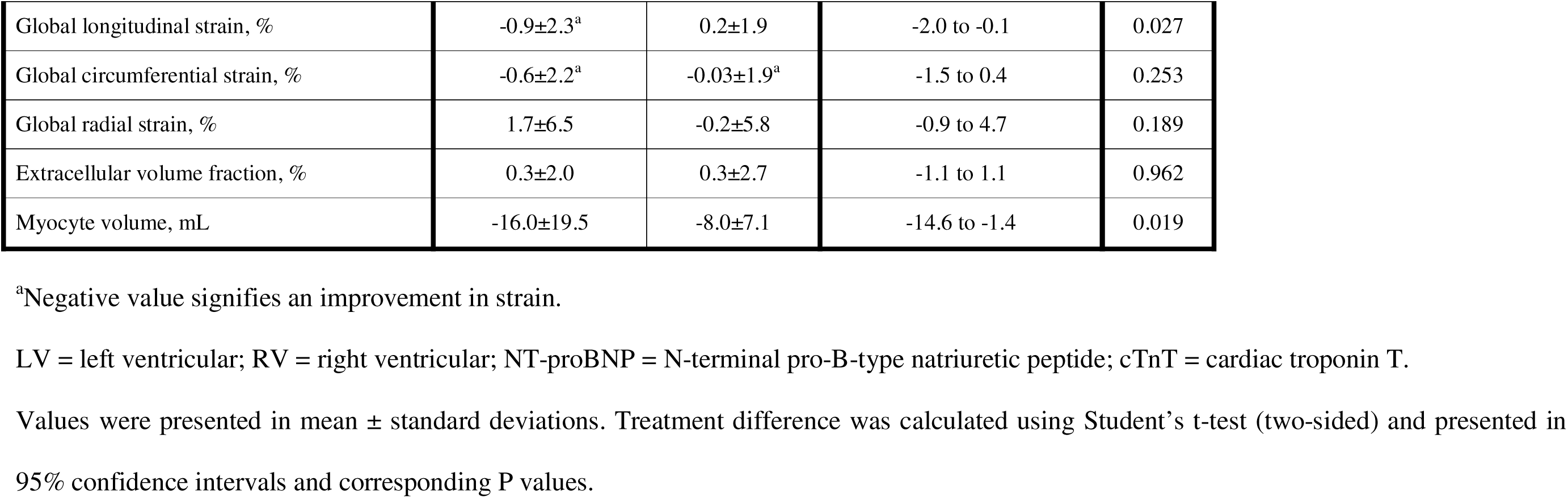
Secondary Endpoints After 52 Weeks of Treatment.

|  | Change from Baseline to Post-treatment |  | Treatment Difference |  |
| --- | --- | --- | --- | --- |
|  | Sacubitril/valsartan | Valsartan | 95% Confidence Interval | P Value |
| LV mass, g | -21.9±25.4 | -11.0±9.0 | -19.5 to -2.3 | 0.014 |
| LV end diastolic volume, mL | -12.4±18.7 | -7.1±16.4 | -13.2 to 2.7 | 0.194 |
| LV end systolic volume, mL | -8.0±18.3 | -5.2±15.7 | -10.5 to 5.0 | 0.482 |
| LV stroke volume, mL | -4.4±9.3 | -1.9±9.4 | -6.7 to 1.7 | 0.243 |
| LV ejection fraction | 1.2±6.5 | 2.1±6.2 | -3.8 to 1.9 | 0.511 |
| RV end diastolic volume, mL | -6.8±11.5 | -4.8±16.5 | -8.4 to 4.5 | 0.545 |
| RV end systolic volume, mL | -2.2±10.5 | -2.4±13.0 | -5.1 to 5.6 | 0.926 |
| RV stroke volume, mL | -4.6±8.6 | -2.4±12.5 | -7.1 to 2.7 | 0.368 |
| RV ejection fraction | -0.3±6.3 | 0.1±8.3 | -3.7 to 2.9 | 0.811 |
| Left atrial volume, mL | -18.3±16.6 | -8.6±13.6 | -16.6 to -2.9 | 0.006 |
| Right atrial area, cm<sup>2</sup> | -1.5±2.5 | -1.3±2.2 | -1.3 to 0.9 | 0.694 |
| LV mass/end-diastolic volume ratio | -0.08±0.09 | -0.04±0.09 | -0.08 to 0.00 | 0.072 |
| Remodeling Index | 0.46±0.56 | 0.38±0.50 | -0.17 to 0.31 | 0.544 |
| Ventricular-arterial coupling ratio | -0.07±0.26 | -0.04±0.23 | -0.14 to 0.09 | 0.655 |
| Estimated LV filling pressure, mmHg | -1.8±1.5 | -0.9±1.1 | -1.5 to -0.3 | 0.003 |
| Maximal wall thickness, mm | -1.1±1.0 | -0.7±0.7 | -0.8 to 0.0 | 0.065 |
| Global longitudinal strain, % | -0.9±2.3<sup>a</sup> | 0.2±1.9 | -2.0 to -0.1 | 0.027 |
| Global circumferential strain, % | -0.6±2.2<sup>a</sup> | -0.03±1.9<sup>a</sup> | -1.5 to 0.4 | 0.253 |
| Global radial strain, % | 1.7±6.5 | -0.2±5.8 | -0.9 to 4.7 | 0.189 |
| Extracellular volume fraction, % | 0.3±2.0 | 0.3±2.7 | -1.1 to 1.1 | 0.962 |
| Myocyte volume, mL | -16.0±19.5 | -8.0±7.1 | -14.6 to -1.4 | 0.019 |

<sup>a</sup>Negative value signifies an improvement in strain.
LV = left ventricular; RV = right ventricular; NT-proBNP = N-terminal pro-B-type natriuretic peptide; cTnT = cardiac troponin T.
Values were presented in mean ± standard deviations. Treatment difference was calculated using Student's t-test (two-sided) and presented in
95% confidence intervals and corresponding P values.

### Treatment Effects on Blood Pressure Control

Participants in both groups achieved similarly lowered 24h mean BP at all timepoints assessed in the trial (Figure 3). At the end of 52 weeks, the 24h mean SBP was 125±11 mmHg with sacubitril/valsartan versus 126±11 mmHg with valsartan (P=0.762 for comparison). This corresponded to a decrease of 13.3±13.1 and 10.6±13.4 mmHg from baseline in the sacubitril/valsartan and valsartan groups, respectively. Treatment difference between the two groups was not significant (-2.7 [95% CI: -8.7 to 3.3] mmHg, P=0.379).

**Figure 3.**
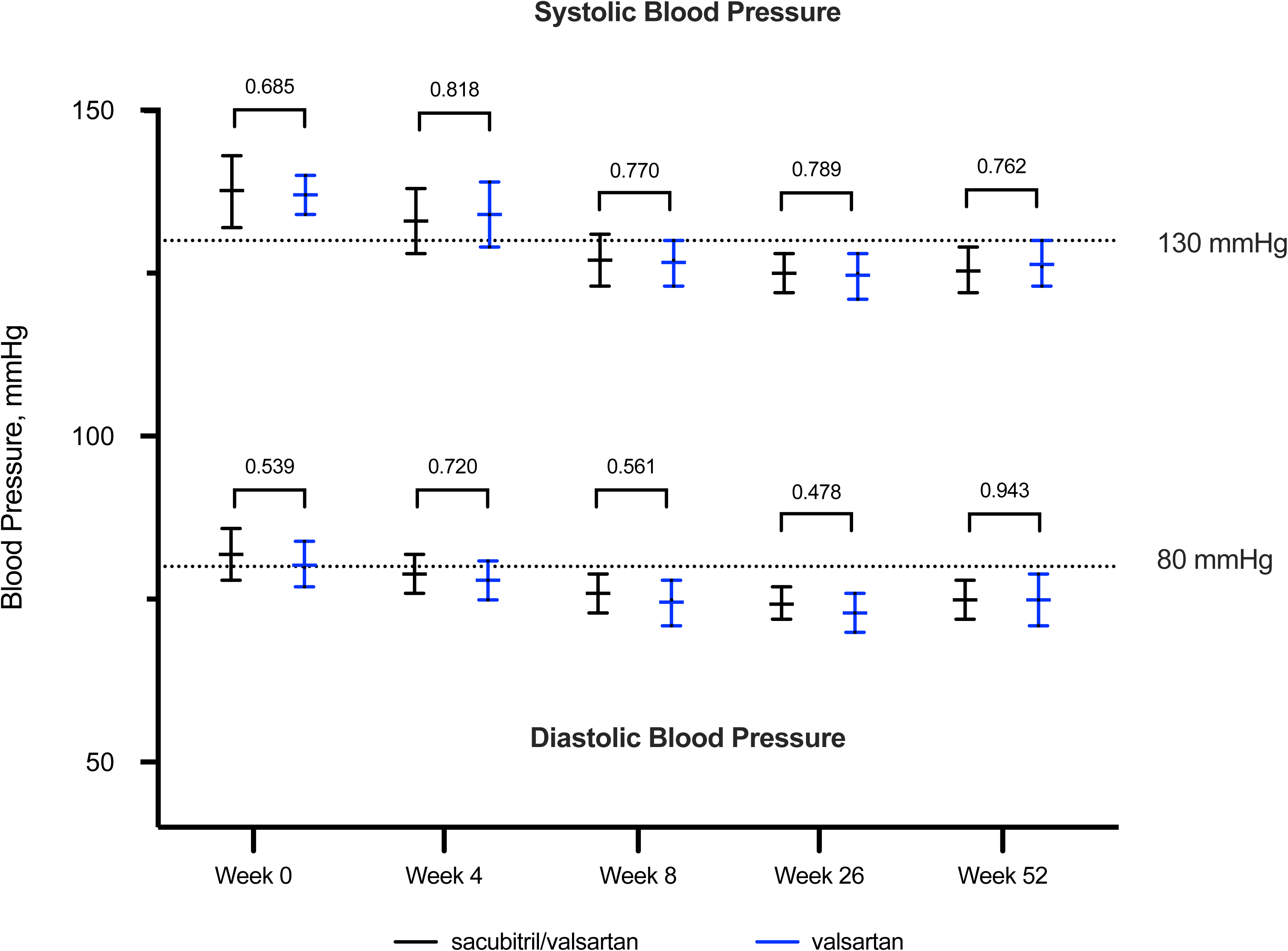
Blood Pressure Control Between Sacubitril/valsartan and Valsartan. The mean blood pressure assessed at all timepoints during the trial were similar between the two treatment groups (n=78). Data were presented in mean and 95% confidence interval; and P values from Student’s t-test (two-sided) comparison.

Similarly, the 24h mean DBP at the end of study was 75±9 mmHg with sacubitril/valsartan versus 75±12 mmHg with valsartan (P=0.943 for comparison), corresponding to a decrease of 7.1±7.1 and 5.6±9.6 mmHg from baseline in the two respective treatment groups. The treatment difference in 24h DBP reduction between the two groups was also not significant (-1.5 [95% CI: -5.4 to 2.3] mmHg, P=0.426).

### Adverse Events

Treatment with sacubitril/valsartan and valsartan were very well-tolerated in this trial. One patient developed nocturnal cough with sacubitril/valsartan three months after randomization. Symptoms improved upon dose reduction, and the participant continued with the trial. Renal function remained stable throughout the trial (Table 3). There were no major adverse cardiovascular events such as hospitalizations for HF, myocardial infarction, strokes and deaths reported during the trial.

**Table 3.** Renal Function Between Treatment Groups Throughout Study Trial.

|  | Week | Sacubitril/<br>valsartan | Valsartan | P value |
| --- | --- | --- | --- | --- |
| Potassium, mmol/L | 0 | 3.8 [3.6, 4.0] | 3.7 [3.6, 4.0] | 0.800 |
|  | 4 | 3.9 [3.7, 4.3] | 4.0 [3.7, 4.3] | 0.778 |
|  | 8 | 4.0 [3.8, 4.2] | 4.0 [3.9, 4.2] | 0.766 |
|  | 26 | 4.1 [3.7, 4.5] | 4.1 [3.9, 4.3] | 0.765 |
|  | 52 | 3.9 [3.6, 4.2] | 3.8 [3.5, 4.2] | 0.602 |
| Estimated GFR,<br>mL/min/1.73m<sup>2</sup> | 0 | 94 [74, 108] | 88 [70, 102] | 0.420 |
|  | 4 | 82 [68, 108] | 84 [74, 100] | 0.996 |
|  | 8 | 86 [71, 102] | 82 [63, 110] | 0.909 |
|  | 26 | 89 [65, 107] | 83 [69, 96] | 0.897 |
|  | 52 | 89 [63, 104] | 83 [69, 100] | 0.796 |

Values were presented in median [inter-quartile range]. Comparisons between treatment groups were performed with Mann-Whitney U (two-sided) test.
GFR = glomerular filtration rate.

## DISCUSSION

The main finding of REVERSE-LVH was that sacubitril/valsartan resulted in a reduction in diffuse interstitial fibrosis in hypertensive patients with LVH following 52 weeks of treatment compared to valsartan alone. This was accompanied by a favorable change in LV mass, circulating cardiac biomarkers and diastolic function (LA size and estimated LV filling pressure). There was no significant difference in systolic function improvement between the two treatment groups.

The use of CMR in REVERSE-LVH has allowed reliable quantification of fibrosis without invasive endomyocardial biopsies, a procedure that was required in previous studies on anti-fibrotic therapies. Novel CMR mapping techniques have dramatically advanced tissue characterization non-invasively, allowing us to investigate novel anti-fibrotic therapies in diverse populations.

ECV fraction represents the ratio of interstitium to the total myocardial volume. Although it is widely used to measure diffuse interstitial fibrosis, its utility may be limited where treatments affect both myocyte and interstitial compartments. In such cases, therapies that reduce both compartments may lead to an unchanged or even increased ECV fraction. For instance, in REVERSE-LVH, despite reductions in interstitial and myocyte volumes, both treatment groups showed a slight increase in ECV fraction, likely due to a greater reduction in myocyte volume relative to interstitial volume. Instead, deeper mechanistic insights can be accomplished by separately assessing the interstitial and myocyte compartments, particularly for novel treatments or interventions with potential effects on both compartments.

Myocardial fibrosis is a hallmark of HF, and not all BP medications have anti-fibrotic properties. Among conventional anti-hypertensive therapies, ACEIs or ARBs that target the RAAS have shown the most promise in regressing biopsy-confirmed myocardial fibrosis in HHD, independent of BP lowering effects. Similarly, mineralocorticoid receptor antagonists (MRAs) like spironolactone and eplerenone reduce serum markers of collagen turnover and improve diastolic function on echocardiography^16–18^.

Sacubitril/valsartan, a first-in-class ARNI used primarily for HFrEF, is also an effective anti-hypertensive medication. While sacubitril/valsartan is approved for hypertension in some countries, its effectiveness in regressing myocardial fibrosis in patients with hypertension has not been established. Through the REVERSE-LVH trial using CMR to assess interstitial volume, sacubitril/valsartan demonstrated a two-fold greater reduction in diffuse interstitial fibrosis compared to valsartan alone. Both treatment groups achieved similar BP throughout the trial, an important feature to isolate the anti-fibrotic effects of sacubitril/valsartan from any potential influence of BP control. It is also noteworthy that the other classes of anti-hypertensive medications prescribed were similar between the two treatment groups.

Sacubitril/valsartan also led to greater LV mass regression compared to valsartan alone, findings that align with results from a previous trial comparing sacubitril/valsartan and olmesartan. Overall, these results reinforced the beneficial impact of sacubitril/valsartan on reverse remodeling in patients with HHD.

PARAGON-HF (Prospective Comparison of ARNI with ARB Global Outcomes in HF with Preserved Ejection Fraction) reported that sacubitril/valsartan did not result in significantly lower rates of HF hospitalizations and cardiovascular deaths in patients with HF and preserved ejection fraction (HFpEF) compared to valsartan^22^. Considering these results, it is important to discuss the relevance of REVERSE-LVH after PARAGON-HF and possible implications for future HFpEF trials.

HFpEF represents a syndrome characterized by phenotypic heterogeneity, underpinned by complex underlying mechanisms, and contributed by various coexisting comorbidities. This heterogeneity is exemplified by the observation that despite similar E/e’ (echocardiographic marker of LV filling pressure), the prevalence of LVH ranges between 14 and 50% in recent HFpEF trials (21% in PARAGON-HF). This observation hints at the presence of additional factors contributing to elevated LV filling pressure (such as chronic lung disease, atrial fibrillation and/or chronic kidney disease). Consequently, it is plausible that not all these patients had derived the anticipated benefits from the HF medications being investigated.

To enhance the effectiveness of future HFpEF trials, one key consideration lies in the strategic inclusion of patients with common phenotypes driving the underlying pathophysiology. This approach will enable more precise selection of pharmaceutical therapies tailored at specific mechanistic phenotypes. For instance, pirfenidone, a small-molecule anti-fibrotic agent without hemodynamic effects, reduced myocardial fibrosis in patients with HFpEF^26^. This reduction in interstitial space without significance changes in myocyte compartment explains for the overall decrease in ECV fraction. In contrast, sacubitril/valsartan reduces both myocardial fibrosis and hypertrophy because of its effects on hemodynamics, natriuretic and RAAS system.

The effectiveness of hypertension treatment is often assessed by the BP targets achieved, with the fundamental goal to minimize end-organ complications such as HF. However, cardiac remodeling in the hypertensive heart is complex and the peripheral BP may not adequately reflect the abnormalities in the myocardium. A moderate correlation was demonstrated between interstitial volume and 24h SBP (r=0.36, P<0.001), worse with office SBP (r=0.21; P<0.01)^27^. This poses a challenge in defining the optimal BP targets in hypertensive patients with LVH, particularly if BP are adequately controlled.

Because the apparent amelioration of diffuse interstitial fibrosis is associated with improved outcomes, the interstitium is an attractive target for drug repurposing and development. Sacubitril/valsartan resulted in a greater reduction in diffuse interstitial fibrosis compared to valsartan alone after 52 weeks of treatment. The observed association with an improvement in diastolic function and circulating biomarkers was encouraging.

The findings from REVERSE-LVH strengthened the case for considering sacubitril/valsartan in selected individuals with hypertensive LVH, especially those who failed to achieve adequate reduction in diffuse interstitial fibrosis with ACEIs or ARBs. Individuals with 24h SBP greater than 135mmHg (median SBP in this study) may have further reduction in diffuse interstitial fibrosis with sacubitril/valsartan but this finding from the post-hoc subgroup analysis was exploratory and should be investigated in future trials.

It was by chance that a higher proportion of female participants (59%) were recruited. Despite the challenges posed by COVID-19 between 2020 and 2021, the trial had very low dropout rates (5%) and exceptionally high treatment adherence rates, surpassing the 80% threshold widely used to define “good adherence”. Both treatment arms achieved similar BP reduction at the end of the trial. These observations highlighted the meticulous conduct of the trial.

The study had some limitations. Two participants (one in each treatment group) were given MRAs, which may confound regression of diffuse interstitial fibrosis. This was inevitable because their BPs were not optimally controlled despite being prescribed the maximum tolerated doses of trial medications and other classes of anti-hypertensives. Although REVERSE-LVH was adequately powered as a phase 2 trial, the effect size observed is small and warrants further investigation in larger studies powered for clinical outcomes. Prior exposure to ACEIs and ARBs in the participants may have introduced confounding, although no significant treatment interactions were observed.

In conclusion, compared to valsartan alone, sacubitril/valsartan appears to exert beneficial myocardial effects beyond blood pressure reduction in patients with hypertensive LVH. However, larger and longer-term studies are necessary to determine the clinical significance of the observed effect sizes.

## METHODS

### Trial Design

The Role of ARNI in the Ventricular Remodeling in Hypertensive LVH (REVERSE-LVH) was a prospective, randomized, open-label, blinded endpoint (PROBE) phase 2 trial (ClinicalTrials.gov identifier: NCT03553810). The original design and methods of the trial has been published^31^ (Supplemental note 1). All study procedures were conducted at the National Heart Centre Singapore (NHCS). This was an investigator-initiated trial. All trial-related expenses, including investigations and medications were funded by the National Medical Research Council. Novartis did not participate in the design of the trial, analysis and interpretation of the data.

REVERSE-LVH was reviewed and approved by the SingHealth Centralized Institutional Review Board (CIRB ref: 2018/2182) and the Health Sciences Authority. All participants provided written informed consent.

### Trial Participants and Recruitment

Participants were recruited between June 2019 and June 2023. These participants were screened and recruited from the REMODEL cohort, an ongoing non-interventional prospective study to examine the role of CMR in patients with hypertension (ClinicalTrials.gov Identifier: NCT02670031). Eligible participants were adults at least 21 years old with essential hypertension and LVH, diagnosed using Asian specific thresholds on CMR^32^.

Essential hypertension was defined as systolic BP (SBP) ≥140 mmHg and/or diastolic BP (DBP) ≥90 mmHg at time of diagnosis, on at least one medication for BP control. Individuals with known secondary causes of hypertension, inherited cardiomyopathies, atrial fibrillation, and history of cardiovascular complications (such as myocardial infarction, HF and stroke) were excluded. Other exclusion criteria included intolerance to ARBs, contraindications for CMR, stage IV or V chronic renal disease, presence of disease with limited life expectancy (<3 years), pregnancy and/or breast-feeding.

### Randomization, Treatment and Blinding

Study participants were randomized to either sacubitril/valsartan or valsartan in a 1:1 fashion. Randomization was performed using sealed and sequentially numbered envelopes, prepared by the Singapore Clinical Research Institute. Crossover of treatment allocation was not allowed, and the treatment duration was 52 weeks. Treatment allocation was not concealed from patients. Treatment assignment and trial endpoints were concealed from the core-lab staff analyzing the CMR images.

Sacubitril/valsartan was initiated at 50-100mg twice a day, uptitrated to maximum 200mg twice a day; valsartan was initiated at 80-160mg once a day, uptitrated to maximum 320 mg once a day. Medications in both treatment groups were uptitrated as tolerated to achieve a target SBP of <140 mmHg according to contemporary guidelines at the time of study conception^33,34^. Additional non-RAAS inhibiting anti-hypertensive agents were prescribed where necessary. Participants who were on ACEI or ARB at the time of enrollment underwent a washout period of two weeks before commencing trial medication to eliminate residual RAAS-intervening effects, during which alternative medicines or uptitration of existing concomitant anti-hypertensive agents were prescribed for BP control.

Study medications were dispensed by NHCS pharmacists after each study visit. Treatment adherence was assessed by pill counting by the pharmacists.

### Study Procedures

Baseline demographic and clinical information were obtained from participants upon randomization. Participants were seen at the clinic with 24h ambulatory BP at weeks 4, 8, 26 and 52. Renal function tests were performed during these visits as part of safety monitoring. Phone calls were made to the participants at weeks 12 and 38. Additional un-scheduled visits were arranged if deemed necessary by the study team (Supplemental Table 1). If withdrawal of participation occurred after 12 weeks of treatment allocation, final visit procedures (including CMR to assess endpoints) would be performed at the time of withdrawal. At each clinic visit, adverse events related to study treatment were assessed by the study team. If present, they were recorded and reported to the local ethics board and the Health Sciences Authority within the required timeline.

Cardiac volumes, function, myocardial mass, and interstitial volumes were assessed at baseline and after 52 weeks of treatment using standardized imaging CMR protocols on the Siemens Aera 1.5T scanner (Siemens Healthineers, Erlangen, Germany). Balanced steady-state free precession cine images were acquired in the long-axis 2-, 3- and 4-chamber views; and short axis cines extending from the mitral valve annulus to the apex (acquired voxel size of 1.6 × 1.3 × 8.0 mm^3^ and 30 phases per cardiac cycle). T1 mapping was performed with the Modified Look-Locker Inversion-recovery sequence, where native T1 maps were acquired using a heartbeat scheme of 5(3)3, and post-contrast T1 maps were acquired 15 minutes after administration of 0.1 mmol/kg gadobutrol (Gadovist; Bayer Pharma AG, Germany) using a heartbeat scheme of 4(1)3(1)2. Details of the imaging protocol were published in the trial protocol^11^.

All CMR images were de-identified and analyzed by trained personnel at the National Heart Research Institute Singapore CMR core-lab using CVI42 software (Circle Cardiovascular Imaging, Calgary, Canada) according to the published analysis protocols^32,35^.

### Primary Trial Endpoint

The primary endpoint was the difference in absolute change in interstitial volume from baseline to post-treatment between the two treatment groups. Interstitial volume, reflecting diffuse interstitial fibrosis in the absence of infiltrative diseases, was derived as the product of ECV fraction × myocardial volume. This approach of quantifying interstitial volume was previously validated against histological fibrosis^10^ and demonstrated prognostic value in patients with HHD^36^.

ECV fraction was measured using the T1 mapping module in CVI42. Hematocrit for calculating ECV fraction was taken on the day of CMR. Myocardial volume (mL) was calculated as myocardial mass (g) divided by the specific density of the myocardium (1.05 g/mL).

### Secondary Trial Endpoints

Secondary endpoints were absolute change in CMR markers of cardiac remodeling that included LV morphology, function (including multi-directional LV strain) and mechanics. LV concentricity was defined by the ratio between LV mass and end-diastolic volume (EDV). Myocyte volume was derived as the product of (1-ECV fraction) and myocardial volume. Maximal left atrial volume was estimated based on the biplane area-length method using the 2- and 4-chamber views, at end atrial diastole^32^.

Several CMR measures of myocardial mechanics were assessed. The Remodeling Index (RI) was used as a marker of LV wall stress ^35,37^. The RI was calculated as 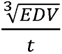 where *t* is the maximal wall thickness (cm) across the 16 myocardial segments. The ventricular-arterial coupling ratio (VCR) was used to describe the relationship between LV and arterial system. It was calculated as the ratio of LV end-systolic volume to stroke volume^38^. LV filling pressures were estimated using a CMR model consisting of left atrial (LA) volume and LV mass, which had been validated with invasively measured pulmonary wedge pressure and demonstrated to have prognostic implications^39^.

Blood samples were collected on the first and final study visit; and stored at −80°C. Biochemical analyses were conducted in a single freeze-thaw cycle across two assay runs at a College of American Pathologists-accredited laboratory (Changi General Hospital, Singapore). Serum NT-proBNP (N-terminal pro-B-type natriuretic peptide; proBNP II STAT, Roche Diagnostics, Germany) and hsTnT (high-sensitivity cardiac troponin T; Trop T hs STAT; Roche Diagnostics, Germany) were measured using electrochemiluminescence immunoassay on the Cobas E801 analyzer (Roche Diagnostics Asia Pacific, Singapore). The lab-reported lower detection limits for NT-proBNP and hsTnT were 11 pg/mL and 3 pg/mL, respectively. All biochemical concentrations below detection thresholds were assigned a value of half the detection limit.

### Sample Size Estimation

An estimated sample size of 70 (35 per treatment group) would provide effective power of 80% at a 5% significance level (two-sided) to detect superiority of sacubitril/valsartan over valsartan for the primary endpoint, assuming an absolute minimum difference of 3.5 mL/m^2^ in mean interstitial volume change between the two groups, a standard deviation (SD) of 5.8mL/m^2^ (data subsequently published^17^) and a moderate correlation of 0.60 between interstitial volume at baseline and week 52. This effect size was based on an estimate of the magnitude of myocardial fibrosis regression that could be expected to translate into improved cardiac function and clinical outcomes^8^. The study plan was to recruit 80 participants, allowing a withdrawal rate of up to 15%.

### Statistical Analysis

The endpoint comparison was based on intention-to-treat analysis, i.e., all participants were analyzed as part of the group to which they had been randomized. Continuous variables were presented as mean ± SD and categorical variables were expressed as count (percentage). Skewed data were presented as medians and inter-quartile range. Comparisons between treatment groups were performed using independent samples t-tests for continuous variables and χ^2^ test for dichotomous variables.

For primary endpoint, analysis-of-covariance (ANCOVA) was performed to adjust for baseline covariates. Treatment group at randomization was included as fixed effect. Baseline interstitial volume, 24h SBP and prior ACEI/ARB use were included as covariates. Results were reported as adjusted mean difference with 95% confidence intervals (CI). Post-hoc subgroup analyses of the primary endpoint were performed using two-way analysis of variance (ANOVA) to test heterogeneity of treatment effects in the following subgroups: age, sex, body mass index, whether washout was performed, LV ejection fraction, interstitial volume and 24h SBP. Continuous covariates were categorized by median values. The hypothesis testing on secondary endpoints was considered exploratory. The analyses were presented with effect estimates and unadjusted 95% CI. Adjustment for multiple comparisons was not performed.

The change in NT-proBNP and hsTnT were reported as the ratio of geometric mean values at baseline and 52 weeks. For treatment comparisons, the effective treatment ratio (ETR) was calculated as the ratio of geometric mean changes in biomarkers between the two groups. An ETR closer to 0 indicates greater effectiveness of sacubitril/valsartan in reducing biomarker level compared to valsartan.

Statistical significance was set at two-sided P value <0.05. All analyses were performed with SPSS, version 24 (SPSS; IBM, INC, Armonk, NY) and R, version 4.3.3 (R foundation for statistical computing, Vienna, Austria). Graphs were created using GraphPad Prism 8.1.2 (GraphPad Software, Inc, San Diego, CA). Statistical analyses were performed by team members who were not involved in image analysis and participants’ follow up.

## DATA AVAILABILITY

De-identified data for this study is available for non-commercial academic purposes. Request for access can be made with the corresponding author and should include a clear research plan with statistical considerations. Requests will be reviewed by the Data Access Committee at the National Heart Centre Singapore within 6 weeks. The data required for the approved, specified purposes will be provided after completion of a data sharing agreement that include instructions on publications, reporting and usage policy. Source data are provided with paper.

## ACKNOWLEDGEMENTS

The authors wish to thank all the study participants, the radiographers at the Department of Cardiovascular Magnetic Resonance, the pharmacists at the Department of Pharmacy and the research coordinators at the Clinical Trial Research Office, National Heart Centre Singapore for their assistance in the study. REVERSE-LVH is funded by the National Medical Research Council of Singapore (MOH-CTGIIT17may-0001).

## AUTHOR CONTRIBUTIONS

CWLC, TTL, SAC, AMR and CHL contributed to the conception and design of the study. QZ designed the statistical analysis plan. VL and DFT led the data collection and management. VL, MD, and QZ performed the data analysis. JAB and TTL led the CMR image analysis. TA analyzed the circulating biomarkers. JB, JD, RF and CSPL provided important inputs to the overall trial conduct. VL and MD wrote the first draft of the manuscript. CWLC secured funding for the trial. All authors made critical revisions for important intellectual content, contributed to the article, and approved the submitted version.

