## Supplementary material for "Effects of Sacubitril/valsartan on Hypertensive Heart Disease: the REVERSE-LVH Randomized Phase 2 Trial": Online Supplemental Data

**Supplemental Information**

Sacubitril/valsartan Compared to Valsartan in Regressing Myocardial Fibrosis in Hypertensive Heart Disease: The REVERSE-LVH Trial

### **Supplemental Table 1. Overview of Study Visits and Procedures**

| **Visit** | **Wash- out** | **1** | **2** | **3** |  | **4** |  | **5** |
| --- | --- | --- | --- | --- | --- | --- | --- | --- |
| **Week** | **-2** | **0** | **4^a^** | **8^a^** | **12^a^** | **26^a^** | **38^a^** | **52** |
| Informed consent taking | X^b^ | X^b^ |  |  | Phone call follow-up |  | Phone call follow-up |  |
| Demographics and anthropometrics collection | X^b^ | X^b^ |  |  |  |  |  |  |
| Collection of past medical history, including medications | X^b^ | X^b^ |  |  |  |  |  |  |
| Urine pregnancy test | X | X | X | X |  | X |  | X |
| Randomization |  | X |  |  |  |  |  |  |
| Health-related quality of life assessment |  | X |  |  |  | X |  | X |
| Study medication dispense |  | X | X | X |  | X |  |  |
| Medication adherence check |  |  | X | X |  | X |  | X |
| Blood collection for renal function |  | X | X | X |  | X |  | X |
| Blood collection for storage |  | X |  |  |  |  |  | X |
| Office blood pressure measurement |  | X | X | X |  | X |  | X |
| 24-hour ambulatory blood pressure monitoring |  | X | X | X |  | X |  | X |
| Cardiovascular magnetic resonance and blood collection for hematocrit |  | X |  |  |  |  |  | X |
| Adverse events evaluation |  | X | X | X |  | X |  | X |

^a^Additional clinic visits are be arranged if necessary.

^b^These procedures were only done once, either at the washout visit or visit 1.

### **Supplemental Table 2. Additional Medications Prescribed**

|  | **Sacubitril/**  **valsartan** | **Valsartan** | **P value** |
| --- | --- | --- | --- |
|  | (n=38) | (n=39) |  |
| **Additional medications, n** | **26** | **24** | 0.571 |
| 1 | 20 | 20 |  |
| 2 or more | 6 | 4 |  |
| **Classes of Medications, n** |  |  | 0.926 |
| Calcium channel blockers | 22 | 20 |  |
| Diuretics (hydrochlorothiazide) | 5 | 4 |  |
| Beta blockers | 2 | 2 |  |
| Others^a^ | 4 | 2 |  |

^a^Includes hydralazine and mineralocorticoid receptor agonist. Comparisons were made using chi-square testing with two-sided P values shown.

### **Supplemental Table 3. Primary Outcome by Subgroups**

|  | **Mean Difference** | **95% Confidence Interval** | **P Value for Interaction** |
| --- | --- | --- | --- |
| **Sex** |  |  | 0.108 |
| Female | -1.1 | -3.6 to 1.4 |  |
| Male | -4.3 | -7.2 to -1.3 |  |
| **Prior ACEI/ARB** |  |  | 0.967 |
| No | -2.8 | -5.5 to -0.2 |  |
| Yes | -2.9 | -6.0 to 0.2 |  |
| **Age, years**^a^ |  |  | 0.464 |
| ≤ 59 | -3.3 | -6.0 to -0.6 |  |
| > 59 | -1.8 | -4.9 to 1.2 |  |
| **LVEF, %**^a^ |  |  | 0.058 |
| ≤ 60 | -1.0 | -3.7 to 1.7 |  |
| > 60 | -4.7 | -7.4 to -2.0 |  |
| **SBP, mmHg**^a^ |  |  | **0.016** |
| ≤ 135 | -0.5 | -3.2 to 2.2 |  |
| > 135 | -5.2 | -8.0 to -2.5 |  |
| **Interstitial volume, mL**^a^ |  |  | 0.231 |
| ≤ 25.3 | -1.3 | -4.0 to 1.4 |  |
| > 25.3 | -2.0 | -4.6 to 0.7 |  |
| **BMI, kg/m^2^**^a^ |  |  | 0.425 |
| ≤ 26.4 | -1.9 | -4.7 to 0.9 |  |
| > 26.4 | -3.5 | -6.3 to -0.7 |  |

Treatment-covariate interaction was tested with two-way ANOVA analysis. The primary endpoint (presented in absolute values, mL) was entered as the dependent variable. Each covariate and randomized treatment arm were entered as fixed factors. ^a^Continuous variables were stratified by median values.

LVEF = Left ventricular ejection fraction; SBP = systolic blood pressure; BMI = body mass index; ACEI = Angiotensin-Converting Enzyme Inhibitor; ARB = Angiotensin Receptor Blocker

### **Supplemental Table 4. Secondary Endpoints^a^ After 52 Weeks of Treatment**

|  | **Sacubitril/**  **valsartan** | **Valsartan** | **P value** |
| --- | --- | --- | --- |
| Indexed LV end diastolic volume, mL/m^2^ | -6.6±8.8 | -4.1±8.6 | 0.219 |
| Indexed LV end systolic volume, mL/m^2^ | -4.1±8.9 | -3.1±8.0 | 0.588 |
| Indexed LV stroke volume, mL/m^2^ | -2.5±5.0 | -1.1±5.6 | 0.247 |
| Indexed LV mass, g/m^2^ | -11.5±10.3 | -6.3±5.2 | **0.007** |
| Indexed RV end diastolic volume, mL/m^2^ | -3.9±6.0 | -2.7±9.6 | 0.490 |
| Indexed RV end systolic volume, mL/m^2^ | -1.2±5.7 | -1.4±7.7 | 0.943 |
| Indexed RV stroke volume, mL/m^2^ | -2.7±4.8 | -1.3±7.3 | 0.330 |
| Indexed left atrial volume, mL/m^2^ | -10.1±8.6 | -5.0±8.0 | **0.009** |
| Indexed right atrial area, cm^2^/m^2^ | -0.8±1.4 | -0.7±1.3 | 0.721 |
| Indexed myocyte volume, mL/m^2^ | -8.4±7.9 | -4.6±4.2 | **0.010** |

^a^Indexed to body surface area**.**

LV = left ventricular; RV = right ventricular.

Data were presented in mean and standard deviations. P values from Student’s t-test (two-sided)

were shown.

### **Supplemental Note 1: Study Protocol**
